# A Rapid SARS-CoV-2 Variant Detection by Molecular-Clamping Based RT-qPCR

**DOI:** 10.1101/2021.04.01.21254484

**Authors:** Shuo Shen, Andrew Y. Fu, Maidar Jamba, Jonathan Li, Mike J. Powell, Aiguo Zhang, Chuanyi M. Lu, Michael Y. Sha

**Affiliations:** DiaCarta Inc., 2600 Hilltop Dr, Richmond, CA 94806.; University of California and VA, Healthcare System, San Francisco, CA 94121.

## Abstract

We applied XNA-based Molecular Clamping Technology to develop a multiplex qPCR assay for rapid and accurate detection of SARS-CoV-2 mutations. A total of 278 previously tested SARS-COV-2 positive samples originating primarily from San Francisco Bay Area were tested, including 139 Samples collected in middle January and 139 samples collected at the end of February 2021, respectively. The SARS-CoV-2 Spike-gene D614G mutation was detected from 58 samples (41.7%) collected in January 2021 and, 78 samples (56.1%) collected in February. Notably, while there were no N501Y mutation detected in samples from January, seven of the February samples were tested positive for the N501Y and D614G mutations. The results suggest a relatively recent and speedy spreading of the UK variant (B.1.1.7) in Northern California. This new Molecular Clamping technology-based multiplex RT-qPCR assay is highly sensitive and specific and can help speed up large scale testing for SARS-CoV-2 variants.

## Introduction

While the worldwide vaccination efforts ongoing, the COVID-19 pandemic is continuing to spread with 120 million cases and 2.7 million deaths to date (March 2021). In the United States alone, there are currently over 30 million cases, and the death toll has passed 550 thousand. With the vaccinations picking up steam and more and more people acquiring immunity to the SARS-CoV-2, the focus is now shifting to more transmissible and potentially vaccine resistant novel variants of the virus. At present, at least four SARS-CoV-2 variants, all present in the USA, the UK B.1.1.7 (501Y V1) [1], South Africa B.1.351 (501Y.V2) [2], Brazil P.1 (501Y.V3) [3] and CAL.20C (20C/S:452R; /B.1.429) [4] [5] variants are of particular concern. Lineage B.1.1.7, is also known as 20I/501Y.V1, variant of concern 20DEC-01 (VOC-20DEC-01, previously written as VOC-202012/01) or commonly as the UK variant. It was found in the southeast of England in early October 2020 and has been observed to be increasing in both Europe and the United States [6–9]. It is estimated to be 40%–80% more transmissible than the wild-type SARS-CoV-2 [10–11].

Looking at the mutations present in these 4 main variants of concern, all of them have the D614G mutation, and three of them share the N501Y mutation (except CAL.20C variant). Both mutations are located on SARS-CoV-2 virus spike protein. With the D614G mutation, the amino acid change from aspartic acid to glycine is caused by an A-to-G nucleotide mutation at position 23403 of the virus genome. This change stabilizes the spike protein and enhances its fitness and infectivity [12]. Similar to D614G, N501Y mutation is also associated with higher virus infectivity and even worse, the N501Y mutation is within the receptor-binding domain (RBD) of the spike protein, featured by stronger binding to ACE2 receptor and significant drop of the original vaccine efficacy [13]. These two mutations, D614G and N501Y, are the two earliest and prominent mutations observed thus far as the COVID-19 pandemic continue to evolve.

Next generation sequencing (NGS) has been the standard method for SARS-CoV-2 variants detection. Although the NGS-based assays could confirm the variants, it is expensive, time consuming and not widely available, limiting its utility in large scale testing demand for SARS-CoV-2 variants detection and monitoring. There has been an urgent need for testing platforms that could detect these variants of concern rapidly and cost-effectively. In this study we applied a Molecular Clamping Technology by using xenonucleic acids (XNA) and developed a multiplex reverse-transcription qPCR assay that can accurately and quickly detects known and emerging SARS-CoV-2 mutations (Fig 1).

**Figure 1.**
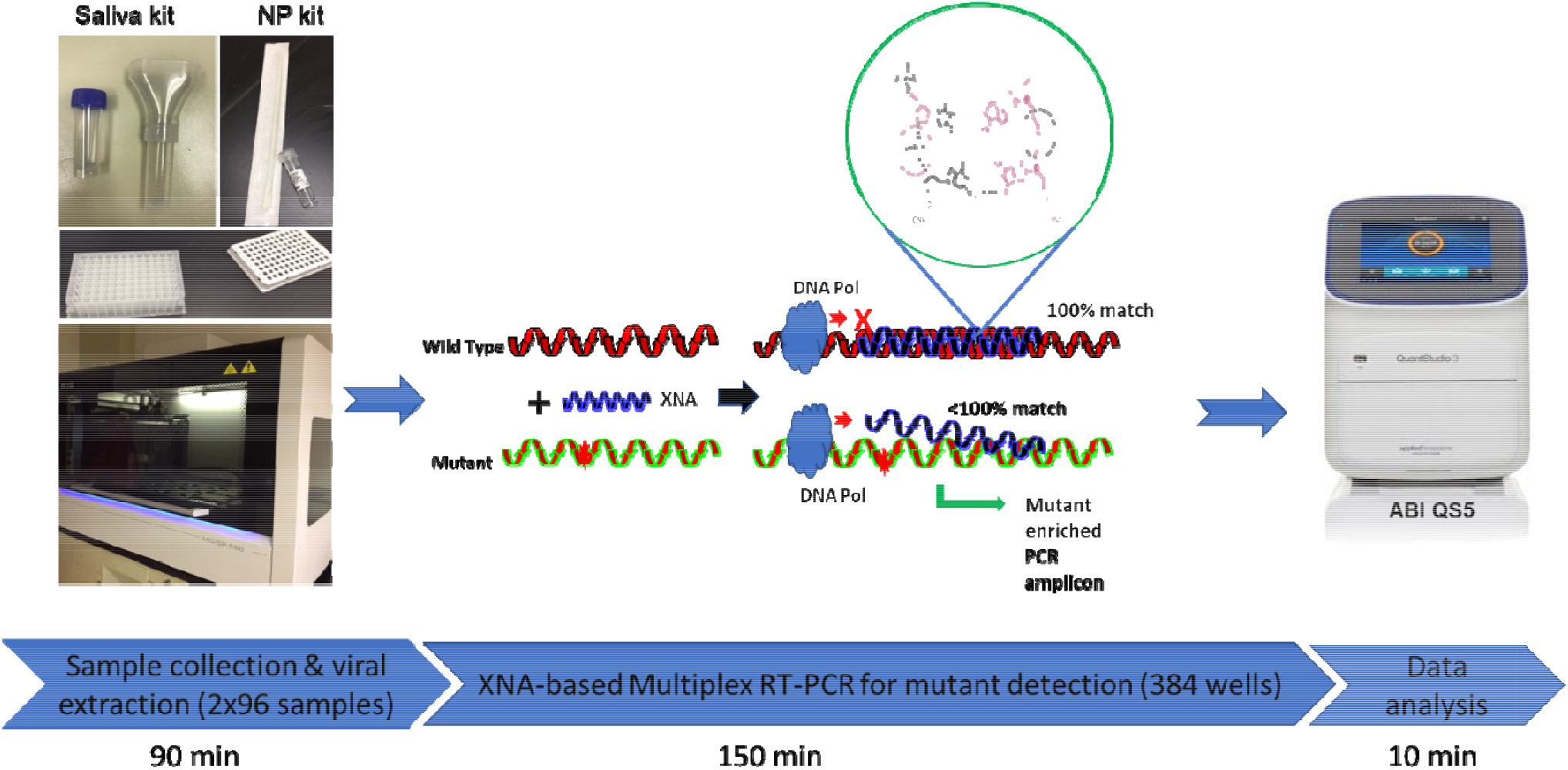
A high-throughput XNA-based Molecular Clomping Technology for SARS-CoV-2 variant detection.

Xenonucleic acids (XNAs) are artificial genetic polymers retaining the Watson-Crick base-pairing capability, originally developed to store genetic information and evolved in response to external stimuli. For practical applications in disease diagnosis and treatment, they can also function as a source of nuclease-resistant affinity reagents (aptamers) and catalysts (xenozymes). More notably, they can be employed as molecular clamps in quantitative real-time polymerase chain reactions (RT-qPCR) or as highly specific molecular probes for detection of nucleic acid target sequences, due to its characteristic and stronger hybridization in XNA/DNA than DNA/DNA [14]. Furthermore, even a single base-pair mismatch between XNA/DNA duplex can result in a drop of 10-18°C in melting temperature Tm [15], allowing the highly specific clamping of XNA molecule onto the targeted sequence (usually wildtype, WT) to block the WT amplification, thus minimizing the WT background in RT-qPCR and selectively enhancing the signal of the mutant. The robust XNAs have been extensively used in the *in vitro* diagnostic assays for detecting cancer-associated gene mutations [16]. Theoretically, XNAs should work in RT-qPCR-based detection of SARS-CoV-2 mutations.

Our report here represents the first attempt of using XNA-based QClamp application in SARS-CoV-2 mutation assay. We expect this fast, reliable and inexpensive mutation detection assay, as we intended to develop, can be made widely available for detecting and monitoring known and emerging SARS-CoV-2 variants during the ongoing CAVID-19 pandemic. Availability and easy access to SARS-Cov-2 mutation testing can also help enable real-time tracking of virus transmission chains and routes in various regions of the world [17].

## Methods

### Study design and ethics

Deidentified leftover patient nasopharyngeal swab (NPS) and saliva samples were used in the study. All patient specimens were collected in January and February 2021 and previously tested at UCSF affiliated San Francisco VA Medical Center clinical laboratory and DiaCarta CLIA certified clinical laboratory for clinical diagnostic or screening purpose. Other than qualitative RT-PCR results (positive or negative), only PCR cycle threshold (Ct) values were included in study analysis and no patient clinical chart reviews were performed. This study was approved by the institutional review board (IRB) at UCSF (UCSF IRB #11-05207) as a no-subject contact study with waiver of consent and as exempt under category 4.

### Sample collection

Saliva and NPS samples of patients were used in the study. All patient specimens were collected, tested and resulted in January and February 2021 and subsequently stored in −80°C freezer. A total of 278 positive samples were selected for this study, including 139 collected in middle January and 139 collected in late February. Other than the qualitative RT-PCR results (positive or negative), only PCR cycle threshold (Ct) values were obtained and included in this study analysis.

### RNA extraction

Automatic RNA/DNA extraction instrument MGISP-960 (MGI Tech Co., Ltd.) and MGI Easy Nucleic Acid Extraction Kit (Cat# 1000020261) was used for the SARS-CoV-2 viral RNA extraction according to the manufacturer’s instructions. Briefly, 180 µL of each nasopharyngeal swab sample or saliva sample was used for extraction. For each batch of clinical samples to be tested, a RNA extraction control (EC) was included (spike 20 µL of EC from the QuantiVirus™ SARS-CoV-2 multiplex kit (DiaCarta, Inc.) into 180 µL sterile RNase-free water). The clinical samples and spiked EC were processed and extracted on the MGI platform. The extraction output is RNA in 30-40 µL RNase-free water, 1 µL of which is used for the RT-qPCR reaction. Precautions were taken while handling extracted RNA samples to avoid RNA degradation. Extracted RNA samples were stored at −80°C if not immediately used for RT-qPCR. The turnaround time from sample extraction to PCR final report is about 4 hrs (Fig 1) [18].

### Multiplex primer and probe design

We targeted the conserved regions of E gene and ORF1ab gene [18] and adjacent areas to the N501Y and D614G mutations in the SARS-Cov-2 genome to design primers and probes for the detection all SARS-CoV-2 variants of concern. Similarly, we designed primers and probes for the human RNase P gene, used as RNA extraction control.

Gene sequences were retrieved from GenBank and GISAID databases for primer and probe design to ensure coverage of all SARS-CoV-2 variant strains. Multiple alignments of the collected sequences were performed using Qiagen CLC Main Workbench 20.0.4., and conserved regions in each target gene were identified using BioEditor 7.2.5. prior to primer and probe designs. Primers and probes were designed to target the most conserved regions of each of the target genes of the viral genome, using Primer3plus software and following general rules of real-time PCR design. All primers were designed with a melting temperature (Tm) of approximately 60 □C and the probes were designed with a Tm of about 65 □C. The amplicon sizes were kept as minimum within the range of 70 bp to 150 bp for each primer pair to achieve better amplification efficiency and detection sensitivity. All designed primers and probes were ordered from Integrated DNA Technologies, Inc. (IDT, Coralville, IA, USA) and LGC Biosearch Technologies (Novato, CA, USA), respectively.

### XNA design, synthesis, purification and analysis

The xenonucleic acids (XNAs) for SARS-COV-2 N501Y and D614G mutations were designed to match the wild type (WT) sequences in order to make them selectively block the qPCR amplification of the WT targets. Each XNA was also designed to partially overlap with and be of the same strand/sense as the corresponding fluorescent probe.

All chemicals and solvents are of ACS grade or higher, purchased from Sigma, Fisher Scientific, Beantown Chemicals, Midland Scientific and other commercial sources. XNAs were synthesized via classic solid-phase peptide synthesis (SPPS) method on an INTAVIS MultiPep automatic synthesizer (INTAVIS Bioanalytical Instruments AG, Cologne, Germany; now a subsidiary of CEM Corporation) [19]. Commercially-available primary Bhoc-protected monomers with aminoethylglycine(AEG)-backbone (Fmoc-“A”, Fmoc-“T”, Fmoc-“C”, Fmoc-“G”), plus terminus-modifying monomers Fmoc-D-Lysine(tBoc) and Fmoc-“O” spacer/linker, were used as the starting materials. TentaGel Resin (from INTAVIS) was chosen and used as the solid support for the multiple-sequence parallel synthesis at a typical 3-umol scale using INTAVIS mini columns. The stepwise SPPS process follows the standard Fmoc chemistry starting from 3’ toward 5’ direction in DMF medium, mainly using HATU for coupling and piperidine for deprotection, within each cycle of solid-phase synthesis, then followed by a new cycle again, keeps repeating itself, untill completion of the entire XNA sequence. After the solid-phase synthesis procedure, the crude product was obtained after a cleavage/deprotection step with a TFA-based cocktail, containing 3-5% triisopropylsilane to minimize side reactions. The resulting off-white crude product then underwent fast purification procedure by size-exclusion chromatography (SEC) with G-25 Sephadex gel (GE Healthcare). For each of 7 XNAs synthesized, right after the SEC step, the highest-concentration fraction, as the main product, was identified by UV quantification at 260 nm (NanoDrop ND-1000) and subsequently analyzed by RP-HPLC (Agilent 1100 HPLC system, Aeris XB-C18 HPLC column of 100 x 4.5 mm, UV detection at 260 nm, column temperature 50 °C), and by mass spectrometry (Shimadzu Axima MALDI-TOF mass spectrometer at UCSF DeGrado Lab). The characteristic XNA identity and molecular weight were confirmed for all 7 XNA products. The selected SEC-fraction of each XNA was thus used for RT-qPCR assay accordingly.

### Optimization of RT-qPCR with XNA

We created three XNAs for N501Y, four XNAs for D614G. A serial dilution XNA qPCR test for N501Y / D614G was conducted to select the best XNA and optimal XNA concentration in the RT-qPCR test.

### Real-time reverse-transcription PCR (rRT-PCR or RT-qPCR)

The XNA-based RT-PCR were set as follows: total volume is 10 ul, including 1.0 µL of RNA, 2.0 µL of primer and probe mixture (final concentration of 0.2 µM and 0.1 µM respectively), 4.5 µL 8 µM of D614G XNA001 and 0.75 µM of N501 XNA003, and 2.5 μL of 4x QuantiVirus SARS-CoV-2 One-step qRT-PCR Master Mix (Catalog# A28526, Thermo Fisher, Waltham, MA). The qPCR was performed at 25 °C for 2 min for uracil-N-glycosylase (UNG) incubation to remove potential carryover, and 53°C for 10 min for reverse transcription, followed by 95 °C for 2 min and then 45 cycles of 95 °C for 3 sec, and 59.5 °C for 30 sec. QuantStudio™ 5 Real-Time PCR System (Thermo Fisher, USA), BioRad CFX384 (Bio-Rad, USA) and Roche LightCycler 480 II (Roche, USA) were used for rRT-PCR amplification and detection [18–20].

### Sanger sequencing verification

All positive samples screened previously by qPCR were sent out for sanger sequencing to confirm their mutational status (SequeTech, CA, USA), and all sequences were analyzed via UCSC SARS-CoV-2 Genome Browser. [21]

### Analysis of the assay sensitivity

The analytical sensitivity of our multiplexed RT-qPCR test with BioRad CFX384 was assessed to give the limit of detection (LoD) data. Since we had confirmed Ct is ~34.28 or 34.24 for ORF1ab or E gene for 4plex assay when the viral concentration about 100 copies/mL [18], we applied this to estimate the assay sensitivity due to no SARS-CoV-2 variant refence is commercially available. We used a two-fold dilution series from 800 copies/mL to 25 copies/mL of the templates in triplicates and confirmed the lowest concentration that was detectable with 95% confidence.

## Results

### XNA design and synthesis

The sequences of XNAs were designed in the context of rational selection of primer/probe for the chosen target gene sequence, intended to match perfectly with the wild-type target DNA sequence. The best sequences (3-4 sequences each target) were iteratively adjusted and selected based on the criterion of multiple major physicochemical factors: sequence length, GC content, purine content and arrangement, self-complementarity, and melting temperature. High overall synthetic yields were found for the D614 XNA group (about 5-10%) while good yields for the N501 XNA group (~5%), translating to a 85-92% single-cycle yield, averagely, across all XNA synthesis here. MALDI-MS Spectra of two representative XNA biomolecules D614 XNA001 and N501 XNA003 were shown in Figure 2 and Supplementary Table 1.

**Table 1.**
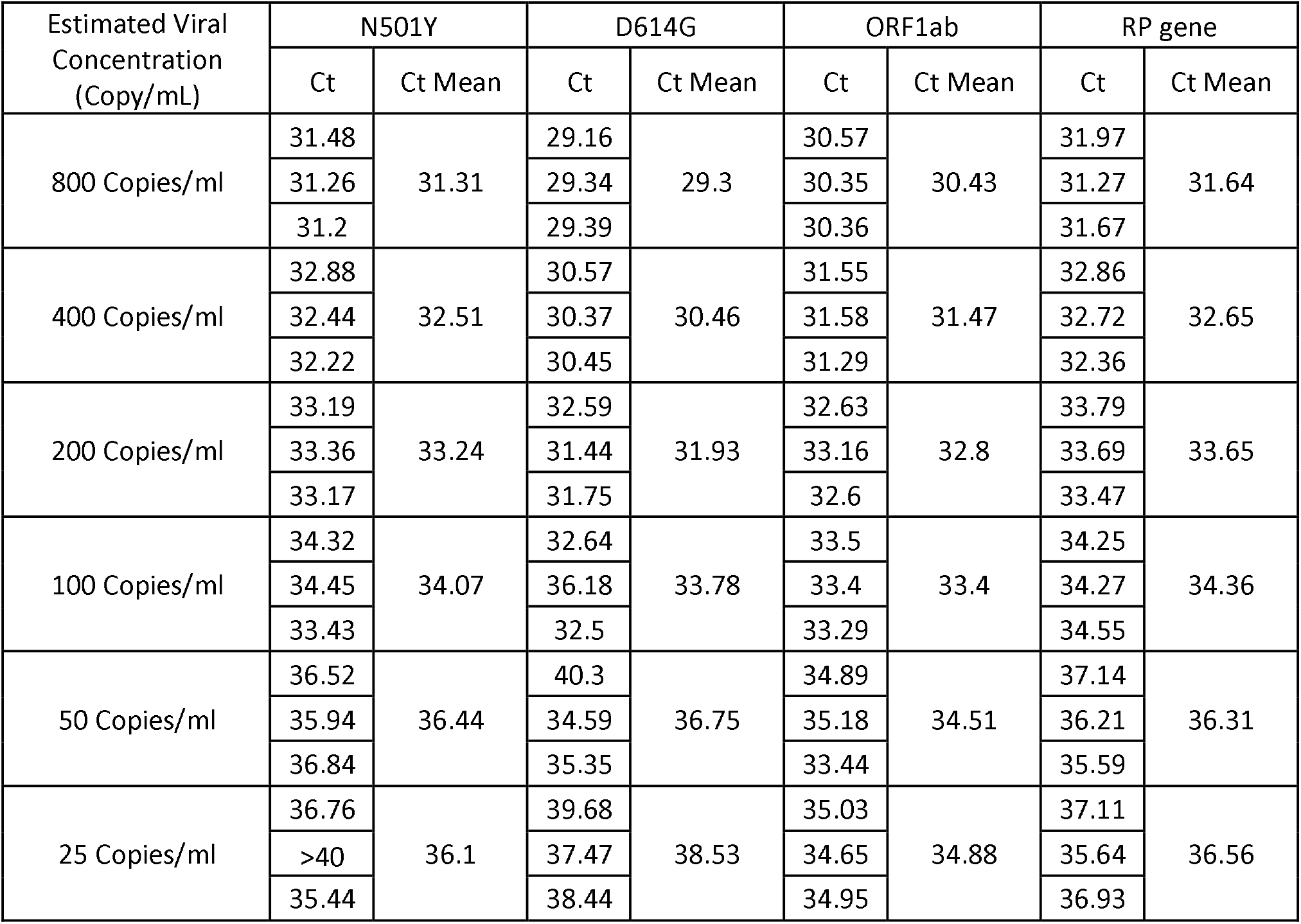
Estimated Limit of Detection (LoD) of the Multiplex RT-qPCR

| Estimated Viral Concentration (Copy/mL) | N501Y |  | D614G |  | ORF1ab |  | RP gene |  |
| --- | --- | --- | --- | --- | --- | --- | --- | --- |
|  | Ct | Ct Mean | Ct | Ct Mean | Ct | Ct Mean | Ct | Ct Mean |
| 800 Copies/ml | 31.48 | 31.31 | 29.16 | 29.3 | 30.57 | 30.43 | 31.97 | 31.64 |
|  | 31.26 |  | 29.34 |  | 30.35 |  | 31.27 |  |
|  | 31.2 |  | 29.39 |  | 30.36 |  | 31.67 |  |
| 400 Copies/ml | 32.88 | 32.51 | 30.57 | 30.46 | 31.55 | 31.47 | 32.86 | 32.65 |
|  | 32.44 |  | 30.37 |  | 31.58 |  | 32.72 |  |
|  | 32.22 |  | 30.45 |  | 31.29 |  | 32.36 |  |
| 200 Copies/ml | 33.19 | 33.24 | 32.59 | 31.93 | 32.63 | 32.8 | 33.79 | 33.65 |
|  | 33.36 |  | 31.44 |  | 33.16 |  | 33.69 |  |
|  | 33.17 |  | 31.75 |  | 32.6 |  | 33.47 |  |
| 100 Copies/ml | 34.32 | 34.07 | 32.64 | 33.78 | 33.5 | 33.4 | 34.25 | 34.36 |
|  | 34.45 |  | 36.18 |  | 33.4 |  | 34.27 |  |
|  | 33.43 |  | 32.5 |  | 33.29 |  | 34.55 |  |
| 50 Copies/ml | 36.52 | 36.44 | 40.3 | 36.75 | 34.89 | 34.51 | 37.14 | 36.31 |
|  | 35.94 |  | 34.59 |  | 35.18 |  | 36.21 |  |
|  | 36.84 |  | 35.35 |  | 33.44 |  | 35.59 |  |
| 25 Copies/ml | 36.76 | 36.1 | 39.68 | 38.53 | 35.03 | 34.88 | 37.11 | 36.56 |
|  | >40 |  | 37.47 |  | 34.65 |  | 35.64 |  |
|  | 35.44 |  | 38.44 |  | 34.95 |  | 36.93 |  |

**Figure 2.**
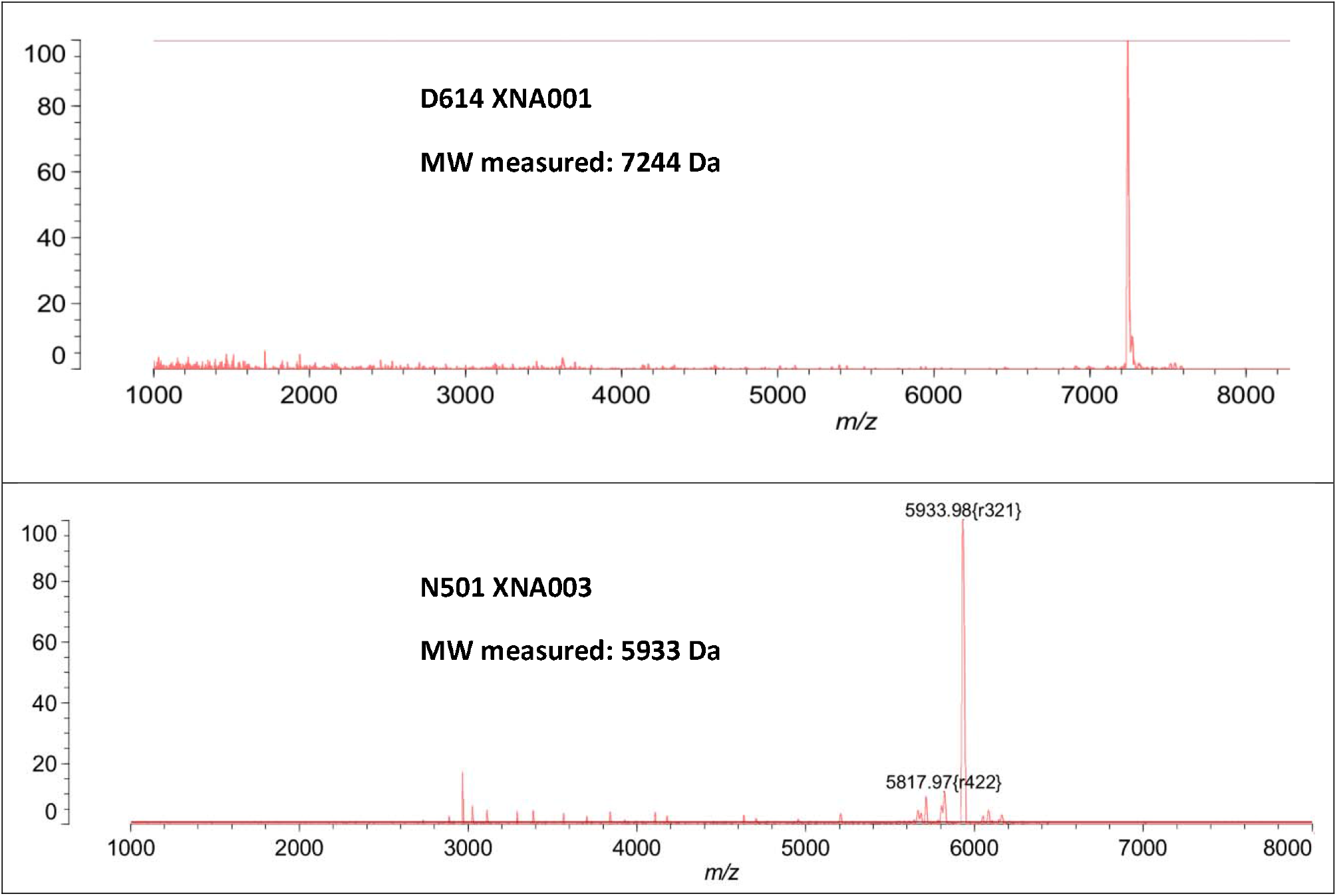
MALDI-MS Spectra of two representative XNA biomolecules: (a) D614 XNA001; (b) N501 XNA003. Molecular weight was determined by the measured cationic (M+H)^+^ peak, for the singly charged parent ion.

### XNA enhance distinguishing mutant to the wild type of SARS-CoV-2

In order to test whether XNA clamping the wild type and enhance mutant detection, we compare a RT-qPCR with and without XNA. An amplification curve of D614G mutant vs wild type without XNA shown that it is difficult to distinguish mutant and wild type (Supplementary Figure 1a). However, with XNA, clearly it is easy to identify mutant to wild type of SARS-CoV-2 (Supplementary Fig 1b). To optimize the assay, we tested different concentration of XNA. The optimization results of XNAs were exemplarily displayed in Supplementary Tables 2 & 3. The XNA that generates the highest delta Ct value between wild type and mutants was selected for the next step in RT-qPCR. Based on the results, we selected D614G XNA 001 & N501Y XNA 003 (Figure 2).

**Table 2.** Summary of Multiplex qPCR testing results of the 278 clinical samples collected in middle January and late February 2021.

| Sample Collection Time | RT-qPCR Test | N501Y | D614G | ORF1ab & E gene* | RNase P gene |
| --- | --- | --- | --- | --- | --- |
| Jan 2021 | Positive | 0 | 58 | 132 | 139 |
|  | Total (N) | 139 | 139 | 139 | 139 |
|  | Detection Rate (%) | 0% | 41.7% | 95.0% | 100% |
| Feb 2021 | Positive | 7 | 78 | 139 | 139 |
|  | Total | 139 | 139 | 139 | 139 |
|  | Detection Rate (%) | 5.0% | 56.1% | 100.0% | 100% |

\*ORF1ab & E gene are targeted for wild type SARS-CoV-2

### Analytical sensitivity

We performed the QClamp multiplex RT-qPCR with XNA on various equipment (BioRad CFX384, Roche Light Cycler and QS5) and all the results were consistent. We diluted the patient sample from 800 copies/mL down to 25 copies/mL and repeated RT-qPCR. Since we had confirmed Ct is ~34.28 for ORF1ab at 4plex SARS-CoV-2 detection assay when the viral concentration about 100 copies/mL [18], we applied this to estimate the assay sensitivity. The data shown that the Ct values were around 33.4 for ORF1ab (wildtype) when estimated viral RNA concentration was at 100 copies/ml. Since N501Y and D614G mutant were detected, and its Ct were 34.07 and 33.78 separately (Table 1), so this indicate that the assay analytical sensitivity limit of detection (LoD) is estimated approximately 100 copies/mL.

### UK variant sped up in bay area of California

In order to test whether there is the UK variant presenting in the San Francisco Bay Area, we screened 139 known SARS-CoV-2 positive samples in January and 139 positive samples in February 2021. Among the 139 positive specimens collected in mid-January 2021, 58 (41.7%) were positive only for D614G but not the N501Y (Table 2). However, among the 139 positive specimens collected at the end of February 2021, there were 7 (5.04%) specimens that were positive for both N501Y and D614G mutations, consistent with the U.K. variant (B1.1.7) (Table 2).

Furthermore, the multiplex RT-qPCR test showed high specificity-amplification of all non-N501Y samples and negative controls were inhibited in the qPCR, while only the N501Y variants and positive controls were amplified (Figure 3).

**Figure 3.**
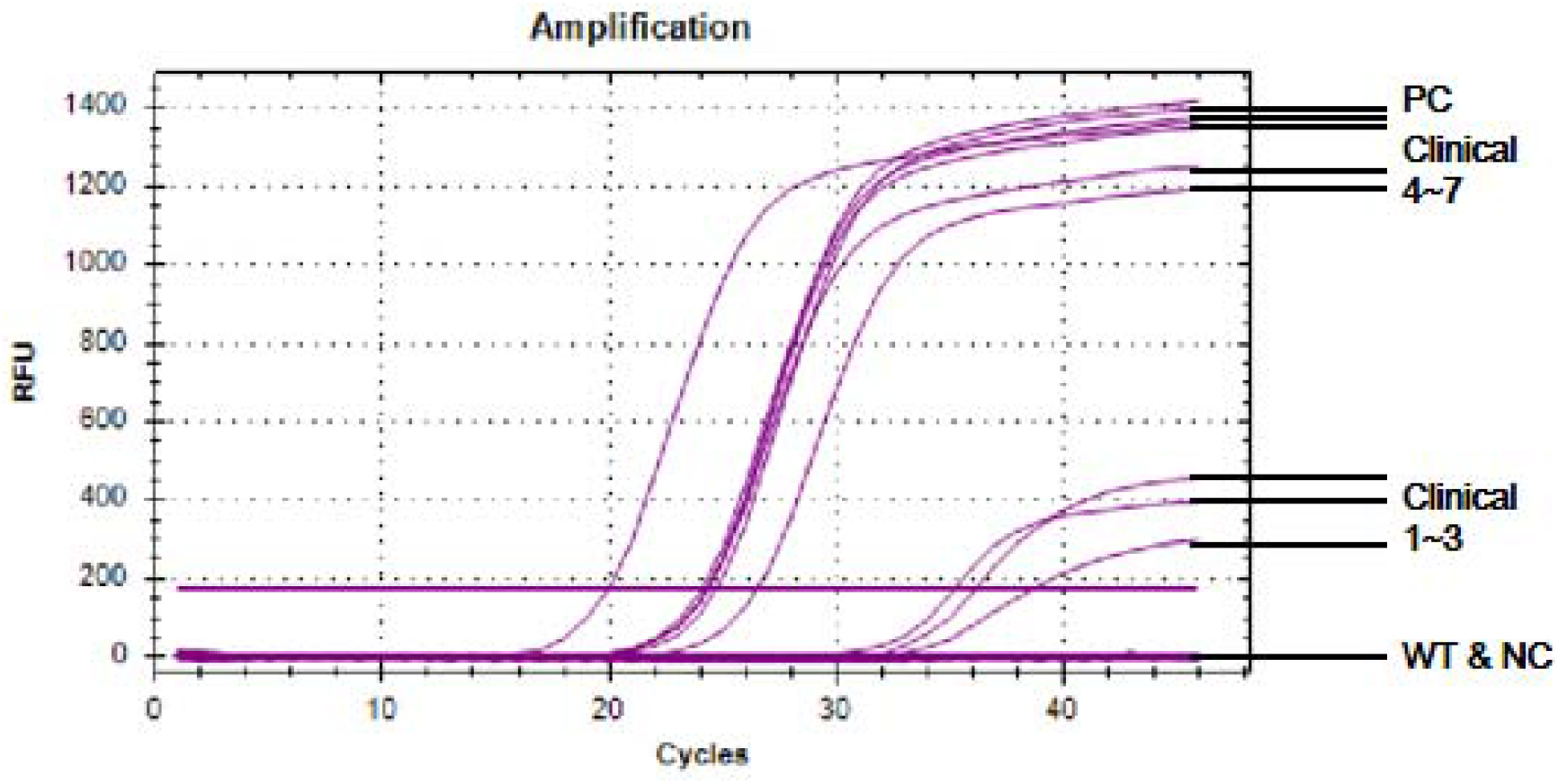
A representative qPCR amplification curve of SARS-CoV-2 variant N501Y. PC, positive control; WT, wild type; NC, negative control.

In order to verify whether these fifty-eight D614G mutant and seven D614G/N501Y mutant samples were truly mutants, we tested these samples’ amplicons by Sanger Sequencing. All of the 58 positive D614G and 7 positive D614G/N501Y mutant samples were confirmed by Sanger Sequencing. The Sanger sequencing peaks showed the target mutant on viral cDNA T <C for D614G (Figure 4a) and the target mutant substitution on viral cDNA A <T for N501Y (Figure 4 b). All 58 positive D614G and 7 positive D614G/N501Y and 213 wild types were confirmed by Sanger sequencing and indicates that its specificity is 100%.

**Figure 4.**
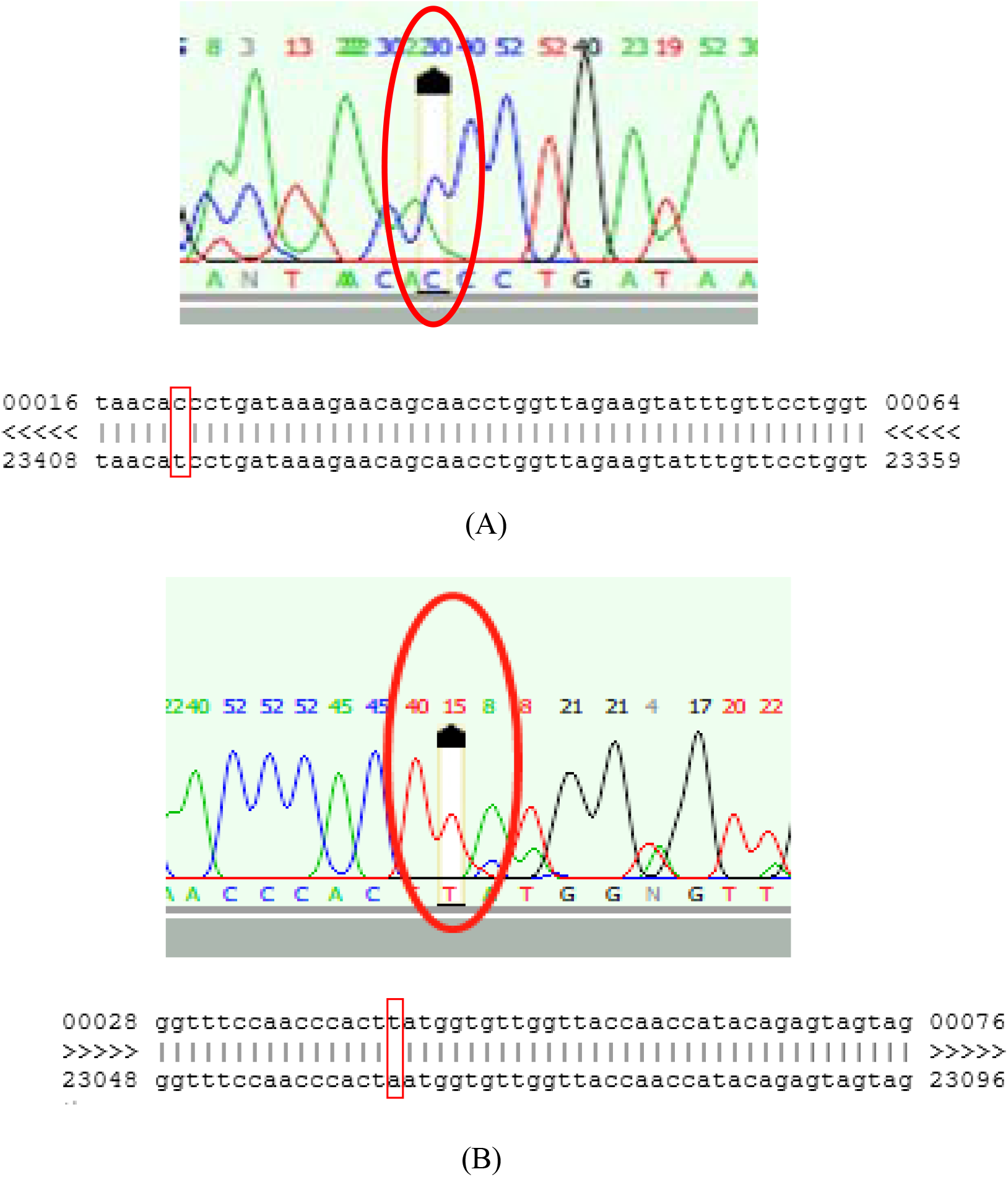
Confirmation of D164G and N501Y mutations by Sanger sequencing. 4a, Sanger sequencing peaks (C in red circle: D614G mutant). Sequence alignment of D614G, Red squares indicate D164G mutant sequence T<C; 4b Sanger sequencing peaks (T in red circle: N501Y mutant). Sequence alignment of N501Y. Red squares indicate N501Y mutant sequence A<T.

## Discussion

For a sensitive and specific molecular clamping QClamp assay, combined with a set of primers and probe for a given gene mutation target, choosing an XNA with appropriate sequence and desired characteristics is important. Our XNA selection process has two folds: first, in the sequence design we already excluded those improper sequences with problematic features (too long or too short, Tm too high or too low, high purine content, purine stretch, unwanted self-complementarity within or between XNA molecules); second, for each mutation assay, 3 or 4 XNAs were designed and synthesized, then compared side-by-side for their performance in RT-qPCR clamping robustness (higher delta-Ct value preferred – greater Ct difference between wildtype gene and housekeeping gene amplification) and clamping specificity (higher delta Ct value preferred, it means greater Ct difference between wildtype gene and mutation gene amplification). Two XNAs, D614 XNA001 and N501 XNA003, were selected respectively for the two mutations detection. Notably, in both groups, the XNA with Tm of nearly 80 °C stands out in selection process, which may be governed by the established RT-qPCR temperature-cycling conditions. [22]

Due to its higher transmissibility the D614G variant is currently the dominant virus variant of SARS-CoV-2 in the United States. Its sub-clade, the N501Y mutation has independently emerged in the UK and South Africa and has been gaining ground recently. Our data suggests that its spread in the Northern California has been a relatively recent event because the patient samples collected in January 2021 were all negative for the N501Y mutation, whereas the N501Y mutation was detected in 5% of the samples collected in late February. This prevalence of B1.1.7 is likely to increase, considering the CDC also forecasted that the N501Y might become the main SARS-CoV-2 variant according to the new epidemiology model [23].

In order to meet the challenge of spreading of SARS-CoV-2 variant strains, availability of rapid and accurate testing platforms is critical. Amongst many detection methods, the qPCR-based methods are regarded as the fastest and most cost-effective, also highly practical in the real-time monitoring of SARS-CoV-2 evolution around the globe. To date, some biotechnology and academic institutions have been developing or published corresponding detection methods using qPCR, and most can be categorized as following: 1) mutant gene specific or allele-specific primer and probe; 2) spike gene target failure (SGTF); 3) E gene target failure and 4) ORF gene deletion [24–27]. Some of them still require NGS to confirm the results. Other testing platforms require pre-testing by regular SARS-CoV-2 RT-PCR tests before variants testing. These issues limit the use of the currently available variant testing methodologies. Of particular note, among these methods, the allele-specific PCR (AS-PCR) quickly attracted attention for mutation detection by some developers. However, AS-PCR has two inherent shortcomings:1) Due to the fixed 3′ end of the allele-specific primer, it is not always feasible to choose optimal primers for PCR amplification; 2) low-quality or cruder DNA/RNA samples could not work similarly well as the good quality ones [28]. In both cases, XNA-based qPCR can help or excel because XNA sequences can be designed with some more flexibility compared to restrictive primer sequence design [29], and because for samples with low-concentration RNA, XNA-based qPCR can realize its full potential in detecting low-concentration, high-background mutant gene targets.

The molecular clamping technology used in this study adds considerably to the sensitivity and specificity to the conventional qPCR. Wild-type background amplification would be minimized. Moreover, multiplexing QClamp qPCR tests could greatly improve the simultaneous detection of multiple targets as well, as we could investigate the wild type of SARS-CoV-2 (ORF1ab gene for wildtype detection) and variants in a single run. Furthermore, in addition to detecting the existing SARS-CoV-2 mutations D614G and N501Y, as demonstrated in this study, the QClamp technology can also be quickly expanded to other emerging mutants such as B.1.351’s E484K, which is of particular concern because it can escape neutralization by vaccine-induced humoral immunity [30]. This strategy can provide an effective and powerful testing platform for known and emerging SARS-CoV-2 variants in the future. This technology can be easily adopted by all clinical laboratories that perform SARS-CoV-2 RT-PCR testing.

CDC descripted 8337 cases of UK variant B1.1.7 and Helix has reported 499 cases of UK variant in California (March 26, 2021) [8–9]. Wetested clinical samples and detected 7 cases of B1.1.7 from 139 randomly selected samples collected in late February 2021. The result suggests that there could be higher prevalence of the UK variant (B1.1.7) in Northern California than previously predicted. Challen et al reported that the mortality hazard ratio associated with VOC-202012/1 (UK variant B.1.1.7) infection was 1.64 in the community and increased in death rate from 2.5 to per 1000 detected case, which translates to a 32-104% increased risk of death. Moreover, its being 40% −80% more transmissible than wildtype SARS-CoV-2, making government agencies and medical communities more worried about this variant [10–11]. The new variant with increased transmissibility would lead to a potentially exponential increase in the resulting number of deaths [10].

In summary, we have developed a multiplex qPCR testing platform for rapid detecting SARS-CoV-2 variant strains, using XNA-based Molecular Clamping Technology. This testing platform can be easily adopted by laboratories that perform SARS-CoV-2 PCR testing, providing a practical solution in lieu of NGS-based testing for detecting and monitoring SARS-CoV-2 variants.

## Data Availability

all data referred to in the manuscript are available to public.

## Acknowledgements

We are grateful for the great help in accessing clinical samples collected from our CLIA Lab colleagues Dr. Yulia Loginova and Eric Abbott. We also, especially chemist A.F., thank UCSF DeGrado research lab for MALDI-MS instrumentation assistance from their group’s Drs. Bobo Dang, Haifan Wu and Hyunil Jo.

## Author Contributions

Data collection, analysis and interpretation (S.S., M. J., A.F., J. L., M.S., C.M.L.); chemical synthesis and analysis (A.F.); clinical sample processing (J. L. and S. S.); writing of original draft (S.S., M. J., A.F., M.S.); revision and editing (M.J., A.F., M.S., S.S., C.M.L, A.Z.); conceptualization (M.S., M. P., A.F., M. J.); project planning and administration (M.S.; A. Z.).

## References

1. Rambaut A et al. A preliminary genomic characterisation of an emergent SARS-CoV-2 lineage in the UK defined by a novel set of spike mutations: COVID-19 genomics UK consortium; December 20, 2020. Available from: https://virological.org/t/preliminary-genomic-characterisation-of-an-emergent-sars-cov-2-lineage-in-the-uk-defined-by-a-novel-set-of-spike-mutations/563.

2. Tegally H et al. Emergence and rapid spread of a new severe acute SARS-CoV-2 lineage with multiple spike mutations in South Africa. medRxiv preprint, posted December 22, 2020. doi: https://doi.org/10.1101/2020.12.21.20248640

3. Faria NR et al. Genomics and epidemiology of a novel SARS-CoV-2 lineage in Manaus, Brazil. medRxiv https://doi.org/10.1101/2021.02.26.21252554; posted March 3, 2021.

4. (a)Zhang W et al. Emergence of a novel SARS-CoV-2 strain in Southern California. medRxiv preprint. Jan 20, 2021. doi: https://doi.org/10.1101/2021.01.18.21249786;(b) Zhang W et al. Emergence of a Novel SARS-CoV-2 Variant in Southern California. JAMA. Feb 11. https://doi.org/10.1001/jama.2021.1612. (2021)

5. Tegally H et al. Emergence of a SARS-CoV-2 Variant of Concern with Mutations in Spike Glycoprotein. Nature. March 1–8. https://doi.org/10.1038/s41586-021-03402-9. (2021)

6. Wise J. Covid-19: New coronavirus variant is identified in UK. BMJ 2020;371:m4857. doi:10.1136/bmj.m4857

7. Risk related to the spread of new SARS-CoV-2 variants of concern in the EU/EEA – first update. Eur. Cent. Dis. Prev. Control. 2021. www.ecdc.europa.eu/en/publications-data/covid-19-risk-assessmentspread-new-variants-concern-eueea-first-update (accessed 30 Jan 2021).

8. CDC. US COVID-19 Cases Caused by Variants. Centers for Disease Control and Prevention. 2020. www.cdc.gov/coronavirus/2019-ncov/transmission/variant-cases.html (accessed 25 March 2021).

9. The Helix COVID-19 Surveillance Dashboard. Helix. www.helix.com/pages/helix-covid-19-surveillance-dashboard (accessed 26 March 2021).

10. Davies et al Estimated transmissibility and impact of SARS-CoV-2 lineage B.1.1.7 in England. medRxiv. doi: https://doi.org/10.1101/2020.12.24.20248822

11. Leung et al. Early transmissibility assessment of the N501Y mutant strains of SARS-CoV-2 in the United Kingdom, October to November 2020. Euro Surveill. 2021;26(1): 2002106. https://doi.org/10.2807/1560-7917.ES.2020.26.1.2002106

12. Korber B et al. Tracking Changes in SARS-CoV-2 Spike: Evidence that D614G Increases Infectivity of the COVID-19 Virus. Cell. 182 (4): 812–827 e819. (2020)

13. Hagen A. SARS-CoV-2 Variants vs. Vaccines. American Society for Microbiology website. https://asm.org/Articles/2021/February/SARS-CoV-2-Variants-vs-Vaccines, mMarch 3, 2021

14. Wang Q et al. Molecular Beacons of Xeno-Nucleic Acid for Detecting Nucleic Acid. Theranostics 3(6): 395–408. doi: 10.7150/thno.5935 (2013)

15. Kyger EM, Krevolin MD and Powell MJ. Detection of the Hereditary Hemochromatosis Gene Mutation by Real-Time Fluorescence Polymerase Chain Reaction and Peptide Nucleic Acid Clamping. Analytical Biochemistry 260: 142–148 (1998)

16. Powell MJ and Zhang A. DNA mutation detection employing enrichment of mutant polynucleotide sequences and minimally invasive sampling. US Patent 1040277B2 (filed in 2015, published in 2016)

17. Bert Ely and Taylor Carter. DISCOVER MAGAZINE article on April 28, 2020. https://www.discovermagazine.com/health/the-coronavirus-genome-is-like-a-shipping-label-that-lets-epidemiologists

18. Sun Q et al. Saliva as a testing specimen with or without pooling for SARS-CoV-2 detection by multiplex RT-PCR test. PLoS ONE 16(2): e0243183. https://doi.org/10.1371/journal. pone. 0243183 (2021)

19. Pinheiro VB and Holliger P. Towards XNA nanotechnology: new materials from synthetic genetic polymers. Trends Biotechnol. 32(6): 321–8 (2014)

20. FDA EUA-approved DiaCarta QuantiVirusTM SARS-CoV-2 Test Kit (Instruction For Use). https://www.fda.gov/media/136809/download (2020)

21. Fernandes JD et al. 2020. “The UCSC SARS-CoV-2 Genome Browser.” Nature Genetics 52 (10): 991–98. https://doi.org/10.1038/s41588-020-0700-8. (2020)

22. Ursula G, Kleider W, Berding C, Geiger A, Ørum H, and Nielsen PE. A Formula for Thermal Stability (Tm) Prediction of PNA/DNA Duplexes. Nucleic Acids Research 26 (21): 5004–6. https://doi.org/10.1093/nar/26.21.5004. (1998)

23. Galloway SE. “Emergence of SARS-CoV-2 B.1.1.7 Lineage United States, December 29, 2020–January 12, 2021.” MMWR. Morbidity and Mortality Weekly Report 70. https://doi.org/10.15585/mmwr.mm7003e2. (2021)

24. Harper H et al. Detecting SARS-CoV-2 variants with SNP genotyping. PLOS ONE February 24, 2021, https://doi.org/10.1371/journal.pone.0243185 (2021)

25. Washington NL et al. Genomic epidemiology identifies emergence and rapid transmission of SARS-CoV-2 B.1.1.7 in the United States. medRxiv 2021.02.06.21251159

26. Artesi M et al. A recurrent mutation at position 26,340 of SARS-CoV-2 is associated with failure of the E-gene qRT-PCR utilised in a commercial dual-target diagnostic assay. Journal of Clinical Microbiology, 58 (10) e01598–20. doi: 10.1128/JCM.01598-20

27. https://www.bioworld.com/articles/503479-covid-19-test-makers-are-adapting-for-variants (2021)

28. Myakishev MV, Khripin Y, Hu S, Hamer DH. High-Throughput SNP Genotyping by Allele-Specific PCR with Universal Energy-Transfer-Labeled Primers. Genome Research. 11: 163–169 doi:10.1101/gr.157901 (2001)

29. https://www.thermofisher.com/us/en/home/life-science/oligonucleotides-primers-probes-genes/custom-dna-oligos/oligo-design-tools.html

30. Wang et al mRNA vaccine-elicited antibodies to SARS-CoV-2 and circulating variants. Nature 2021 https://doi.org/10.1038/s41586-021-03324-6

