## Supplemental for "A Rapid SARS-CoV-2 Variant Detection by Molecular-Clamping Based RT-qPCR"

S. Table 1. Matrix-assisted laser desorption/ionization mass spectrometry (MALDI-MS) confirmation of the structure and molecular weights (MW) of 7 XNAs synthesized by SPPS method.

| **XNA Code** | **XNA Length** | **Theoretical MW (Dalton)** | **Measured MW (Dalton)** |
| --- | --- | --- | --- |
| D614 XNA001 | 25-mer | 7245 | 7244 |
| D614 XNA002 | 22-mer | 6453 | 6451 |
| D614 XNA003 | 20-mer | 5914 | 5910 |
| D614 XNA004 | 18-mer | 5372 | 5376 |
| N501 XNA001 | 17-mer | 5097 | 5105 |
| N501 XNA002 | 19-mer | 5599 | 5599 |
| N501 XNA003 | 20-mer | 5936 | 5933 |
| Bovine Cytochrome C  (MW Standard) | N/A | 12232 | 12228 |

S Table 2 Summary of D614G XNA evaluation and selection

| XNA Concentration (uM) | XNA001 | | | | | XNA002 | | | | |
| --- | --- | --- | --- | --- | --- | --- | --- | --- | --- | --- |
|  | Wild Type | | Mutant | |  | Wild Type | | Mutant | |  |
|  | Ct | Ct mean | Ct | Ct mean | Δ Ct | Ct | Ct mean | Ct | Ct mean | Δ Ct |
| 8uM | 50.0 | 50.0 | 33.7 | 34.1 | 15.9 | 31.6 | 31.3 | 30.9 | 30.7 | 0.6 |
|  | 50.0 |  | 34.5 |  |  | 30.9 |  | 30.7 |  |  |
|  | 50.0 |  | 34.0 |  |  | 31.3 |  | 30.5 |  |  |
| 4uM | 50.0 | 41.2 | 30.9 | 30.9 | 10.3 | 31.1 | 30.8 | 30.3 | 30.3 | 0.5 |
|  | 36.5 |  | 30.9 |  |  | 30.7 |  | 30.3 |  |  |
|  | 37.2 |  | 30.8 |  |  | 30.5 |  | 30.3 |  |  |
| 2uM | 36.6 | 36.5 | 30.3 | 30.8 | 5.6 | 30.8 | 30.5 | 30.1 | 32.5 | -2.0 |
|  | 36.2 |  | 31.9 |  |  | 30.4 |  | 37.3 |  |  |
|  | 36.6 |  | 30.3 |  |  | 30.3 |  | 29.9 |  |  |
| 1uM | 36.5 | 36.2 | 30.1 | 30.0 | 6.3 | 30.5 | 30.7 | 29.9 | 30.0 | 0.7 |
|  | 36.5 |  | 30.0 |  |  | 30.7 |  | 30.2 |  |  |
|  | 35.7 |  | 29.8 |  |  | 30.9 |  | 29.9 |  |  |
| 0uM | 29.4 | 29.3 | 30.0 | 30.2 | -0.8 | 30.2 | 30.6 | 30.6 | 31.3 | -0.7 |
|  | 29.0 |  | 30.0 |  |  | 30.3 |  | 31.6 |  |  |
|  | 29.5 |  | 30.5 |  |  | 31.2 |  | 31.6 |  |  |

S. Table 3. Summary of N501Y XNA evaluation and selection

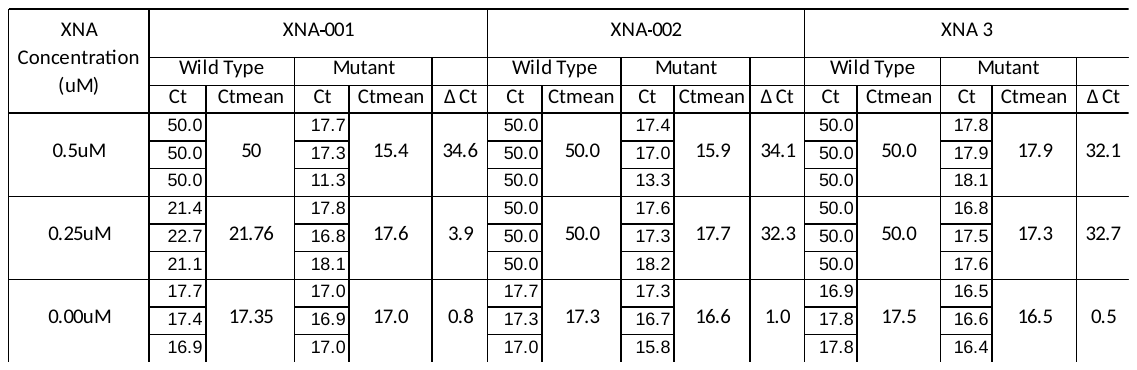

(A)

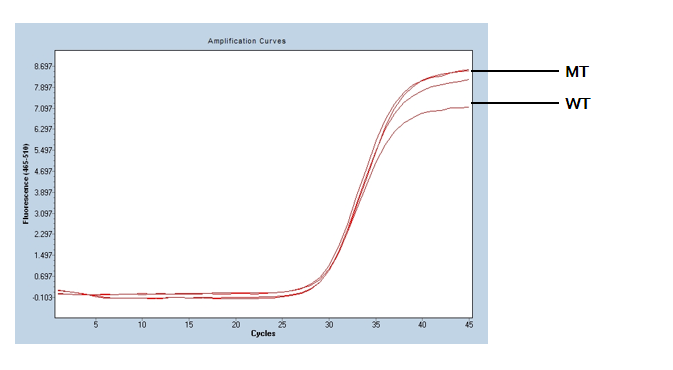

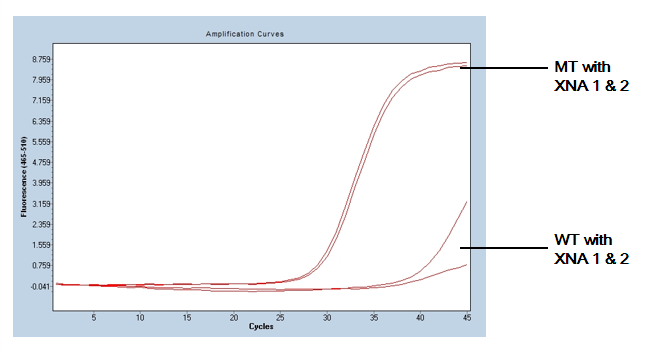

(B)

S. Figure 1. qPCR tests with and without XNA. 1a. Amplification curve of D614G mutant (MT) vs wild type (WT) without XNA. Both wild-type and mutant had ~30 Ct. It is difficult to distinguish mutant and wild type. 1b. Amplification curve of D614G mutant vs wild type with XNA. Wild type amplification was blocked and had high Ct ~40, but mutant was enhanced and had Ct ~27, making the distinction apparent and interpretation intuitive.

(A)

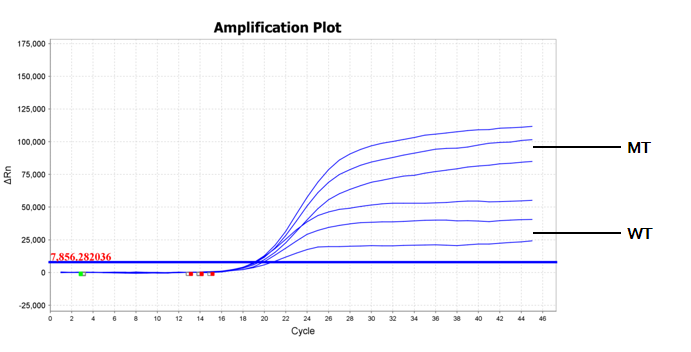

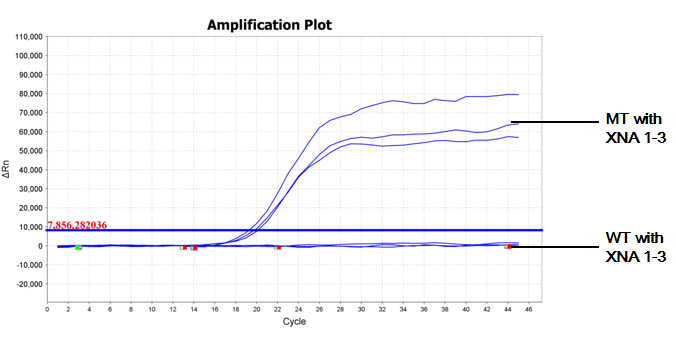

(B)

S. Figure 2. qPCR test with and without XNA. 2a. N501Y mutant (MT) vs wild-type (WT) without XNA. Both wild-type and mutant had ~21 Ct. It is difficult to distinguish mutant and wild type. 2b. N501Y mutant vs wild type with XNA. Wild-type amplification was blocked and had high Ct >40, but mutant was enhanced and had Ct ~20, making the distinction apparent and interpretation intuitive.
